# Sleep macro-architecture and dementia risk in adults: Meta-analysis of 5 cohorts from the Sleep and Dementia Consortium

**DOI:** 10.1101/2024.11.05.24316677

**Authors:** Stephanie Yiallourou, Andree-Ann Baril, Crystal Wiedner, Jeffrey R. Misialek, Christopher E. Kline, Stephanie Harrison, Ethan Cannon, Qiong Yang, Rebecca Bernal, Alycia Bisson, Dibya Himali, Marina Cavuoto, Antoine Weihs, Alexa Beiser, Rebecca F. Gottesman, Yue Leng, Oscar Lopez, Pamela L. Lutsey, Shaun M. Purcell, Susan Redline, Sudha Seshadri, Katie L. Stone, Kristine Yaffe, Sonia Ancoli-Israel, Qian Xiao, Eleni Okeanis Vaou, Jayandra J. Himali, Matthew P. Pase

## Abstract

**Study objectives:** Poor sleep may play a role in the risk of dementia. However, few studies have investigated the association between polysomnography (PSG)-derived sleep architecture and dementia incidence. We examined the relationship between sleep macro-architecture and dementia incidence across five US-based cohort studies from the Sleep and Dementia Consortium (SDC).

**Methods:** Percent of time spent in stages of sleep (N1, N2, N3, REM sleep), wake after sleep onset and sleep maintenance efficiency were derived from a single night home-based PSG. Dementia was ascertained in each cohort using its cohort-specific criteria. Each cohort performed Cox proportional hazard regressions for each sleep exposure and incident dementia, adjusting for age, sex, body mass index, anti-depressant use, sedative use, and *APOE e4* status. Results were then pooled in random effects meta-analyses.

**Results:** The pooled sample comprised 4,657 participants (30% women) aged ≥60 years (mean age was 74 years at sleep assessment). There were 998 (21.4%) dementia cases (median follow-up time of 5 to 19 years). Pooled effects of the five cohorts showed no association between sleep architecture and incident dementia. When meta-analyses were restricted to the three cohorts which had dementia case ascertainment based on DSM-IV/V criteria (n=2,374), higher N3% was marginally associated with an increased risk of dementia (HR: 1.06; 95%CI: 1.00-1.12, per percent increase N3, p=0.050).

**Conclusions:** There were no consistent associations between sleep macro-architecture measured and the risk of incident dementia. Implementing more nuanced sleep metrics remains an important next step for uncovering more about sleep-dementia associations.

**STATEMENT OF SIGNIFICANCE:** Poor sleep may represent a potential lifestyle risk factor for dementia. Sleep is thought to be important for the clearance of toxic Alzheimer’s disease proteins, but whether sleep is associated with dementia risk remains unclear. In the largest study of its kind, utilizing overnight polysomnographic assessment of sleep and data from 5 large U.S cohort studies, we examined the association between sleep macro-architecture and dementia risk. Meta-analysis revealed no clear associations between sleep measures and dementia risk, though there was a suggestion that a higher proportion of N3 sleep may be associated with greater dementia risk. Further exploration of sleep patterns across time, latent sleep traits across metrics, and sleep micro-architecture remain as important next steps for understanding sleep-dementia associations.

## INTRODUCTION

Sleep undergoes significant changes with aging. As adults age, sleep can become less restorative with alterations in sleep architecture. Specifically, there is a shift towards spending more time in lighter sleep stages (N1 and N2), while the duration of slow wave sleep (N3) and rapid eye movement (REM) sleep diminish^1^. Slow wave sleep is thought to affect synaptic plasticity^2^. It also appears to facilitate the clearance of Alzheimer disease proteins^3–5^, though recent findings have challenged this notion^6^. Animal studies indicate that glymphatic clearance of amyloid-β, involved in the formation of amyloid-β plaques in Alzheimer disease, is highest during slow wave sleep^7^. Due to this and other mechanisms, sleep dysfunction may serve as a potential target to reduce the risk of, or delay the onset of dementia^8^. However, in humans, the relationship between sleep and dementia is proving to be complex, as sleep disturbances are a common feature of dementia and the temporal association between sleep architecture and dementia onset remains unclear.

Results derived from studies with an objective sleep assessment, including gold standard polysomnography (PSG), accompanied by long-term follow-up of dementia are limited^9, 10^. To address this gap, we recently formed the Sleep and Dementia Consortium (SDC), comprising five US community-based cohort studies with PSG and long-term follow-up of cognitive, brain imaging and dementia outcomes^11^. Utilizing SDC data (N=5946, mean baseline ages of 58 to 89 years across cohorts), we recently found little evidence of consistent associations between sleep stage percentages and cognitive performance within the next five years; however, sleep disruption measures (poorer sleep efficiency and higher wake after sleep onset, WASO) and the presence of mild to severe obstructive sleep apnea (OSA) were associated with worse global cognition^11^. Other studies have also demonstrated similar associations between sleep disruption and cognition^12–14^. However, whether differences in sleep architecture are associated with dementia incidence remains equivocal with further studies needed. Accordingly, we aimed to examine the association between sleep architecture, measured with a single overnight PSG, and incident all-cause dementia in five community-based cohort studies from the SDC.

## METHODS

The SDC has been described previously.^11^ Briefly, it consists of five community-based cohorts that have performed methodologically consistent, overnight, home-based PSG, as well as cognitive testing and dementia case ascertainment. The cohorts include the Atherosclerosis Risk in Communities (ARIC) study, Cardiovascular Health Study (CHS), Framingham Heart Study (FHS), Osteoporotic Fractures in Men Study (MrOS), and Study of Osteoporotic Fractures (SOF). Written informed consent was provided by all participants prior to the commencement of the study. The study was approved by the Monash University Human Research Ethics Committee, and each cohort obtained institutional review board approval at their respective institutions. For CHS and ARIC, analyses were limited to those with available DNA who consented to genetic studies. Study method and results are reported following the Strengthening the Reporting of Observational Studies in Epidemiology (STROBE) Statement for cross-sectional studies^15^.

### Participants

A brief description of the participating cohorts is provided in the Supplementary Methods. We included participants at least 60 years old who were free of dementia and other neurological disorders (e.g., stroke, multiple sclerosis, significant head trauma, subdural hematoma, brain tumor) at the time of the sleep study and who had PSG data and information on dementia status available during follow-up. Participants with less than 180 minutes of total sleep time or less than one minute of REM sleep were excluded to avoid potential bias from spurious data. Supplementary Table 1 shows the sample selection across cohorts.

### Sleep Metrics

Sleep was measured at baseline. All cohorts used a standardized protocol to complete overnight home-based Type II PSG using Compumedics PSG equipment (Abbotsford, Australia); the model varied by cohort^16–18^). Briefly, EEG (C3-A2 and C4-A1), electrooculogram, electromyogram, thoracic and abdominal displacement (inductive plethysmography bands), airflow (nasal-oral thermocouples and nasal pressure [in MrOS, SOF]), finger pulse oximeter, a single bipolar electrocardiogram, body position by a mercury gauge sensor, and ambient light level were all recorded. Details of the montages for each study are provided at the National Sleep Research Resource (sleepdata.org).

All sleep variables were calculated centrally to ensure consistency of analysis and effective harmonization. Respiratory metrics were annotated and sleep was initially scored in 30-second epochs according to established guidelines (Rechtschaffen and Kales & American Sleep Disorders Association arousal criteria)^17, 18^. The following sleep metrics were calculated: Stage 1 (N1%), Stage 2 (N2%), Stage 3 (N3%), REM sleep (REM%), WASO (total minutes spent awake between sleep onset and offset), sleep maintenance efficiency (SME%) (total sleep time / sleep period time [the time between sleep onset and sleep offset]), total sleep time (minutes), and the apnea-hypopnea index (AHI) (defined as the number of obstructive apneas plus the number of hypopneas accompanied by a greater than 30% reduction in airflow and 4% or greater oxygen desaturation or arousal per hour of sleep). Sleep recording in the ARIC, CHS, and FHS cohorts were limited by a maximum battery life to 9 hours, which prevented further examination of sleep duration greater than 9 hours across all cohorts. Therefore, sleep duration was expressed as ≤6 hours vs > 6 hours (reference). Sleep stages were expressed as a percentage of total sleep duration. Scoring was performed at the time of data acquisition by centrally trained and certified polysomnologists with documented high levels of inter- and intrascorer reliability, as described previously^17^. As some sleep metrics were not normally distributed, square root transformation was applied to N1% and N3% and natural log transformation was applied to SME%, WASO and the AHI.

### Covariates

Co-variates were chosen based on prior knowledge of confounding variables in the association between sleep and dementia. The following co-variates were assessed at baseline and included in statistical models: age (years), sex (men vs. women), body mass index (kg/m2), anti-depressant use (yes vs. no), sedative use (yes vs. no), and APOE *e4* status (non-*e4* carrier vs. at least one copy of *e4*).

### Dementia Case Ascertainment

Dementia case ascertainment is described in detail for each cohort in the Supplementary Methods. Briefly, ARIC, CHS, FHS, and SOF adjudicated dementia diagnosis via varying combinations of neurocognitive data, informant interview, hospitalization records, and based broadly on the Diagnostic and Statistical Manual of Mental Disorders, 4^th^ or 5^th^ edition (DSM-IV/V) criteria. In addition, MrOS investigators adjudicated clinically significant cognitive impairment by a report of physician-diagnosed dementia, use of dementia medication, or a change in modified Mini-Mental State Examination scores ≥ 1.5 standard deviations worse than the mean change from baseline to any follow-up visit. ARIC, CHS and FHS all had continuous surveillance of dementia with dementia adjudicated by a committee according to DSM-IV/V criteria (or equivalent). For CHS, continuous surveillance was performed through to 1998-99, and dementia cases were identified via multiple data sources thereafter (e.g., medications and ICD-9 codes). Both MrOS and SOF assessed dementia at discrete follow-up time points (e.g., several years), up to approximately 10- and 5-years following sleep assessment, respectively.

#### Statistical methods

Statistical analysis was performed using SAS version 9.4 (SAS Institute Inc., Cary, NC, USA) and R version 4.3.0 (R Foundation for Statistical Computing, Vienna, Austria). R code for meta-analysis is made available in the supplementary methods. Demographic characteristics were examined by study cohort. Cox proportional hazards regression models were used to examine the association between sleep metrics and incident all-cause dementia by estimating hazard ratios (HRs) with 95% confidence intervals (CIs). Follow-up duration ranges for each cohort are provided in the Supplementary Methods, and follow-up median durations are provided in Table 1. For each cohort, dementia follow-up commenced from the date of the PSG study to the event of dementia. Non-events were censored at death or until the last date they were known to be dementia-free, or until administrative censoring. Statistical models were adjusted for co-variates listed above. The proportional hazard assumption was examined by including an interaction term between the sleep exposure and the log of follow-up time. The assumption was confirmed graphically and statistically (p-value>0.05) in all cohorts.

**Table 1:**
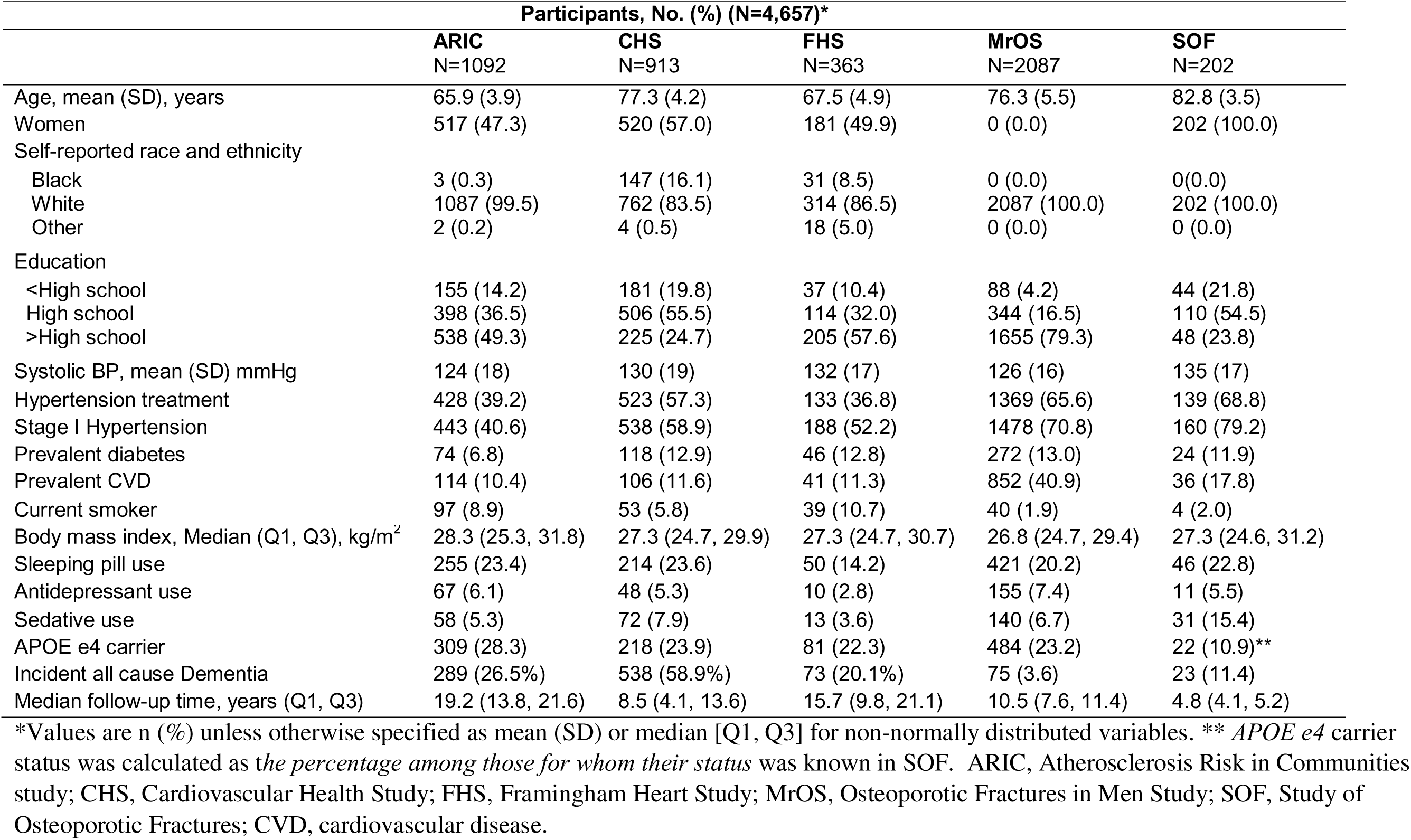
Demographic characteristics.

|  | Participants, No. (%) (N=4,657)* |  |  |  |  |
| --- | --- | --- | --- | --- | --- |
|  | ARIC<br>N=1092 | CHS<br>N=913 | FHS<br>N=363 | MrOS<br>N=2087 | SOF<br>N=202 |
| Age, mean (SD), years | 65.9 (3.9) | 77.3 (4.2) | 67.5 (4.9) | 76.3 (5.5) | 82.8 (3.5) |
| Women | 517 (47.3) | 520 (57.0) | 181 (49.9) | 0 (0.0) | 202 (100.0) |
| Self-reported race and ethnicity |  |  |  |  |  |
| Black | 3 (0.3) | 147 (16.1) | 31 (8.5) | 0 (0.0) | 0(0.0) |
| White | 1087 (99.5) | 762 (83.5) | 314 (86.5) | 2087 (100.0) | 202 (100.0) |
| Other | 2 (0.2) | 4 (0.5) | 18 (5.0) | 0 (0.0) | 0 (0.0) |
| Education |  |  |  |  |  |
| <High school | 155 (14.2) | 181 (19.8) | 37 (10.4) | 88 (4.2) | 44 (21.8) |
| High school | 398 (36.5) | 506 (55.5) | 114 (32.0) | 344 (16.5) | 110 (54.5) |
| >High school | 538 (49.3) | 225 (24.7) | 205 (57.6) | 1655 (79.3) | 48 (23.8) |
| Systolic BP, mean (SD) mmHg | 124 (18) | 130 (19) | 132 (17) | 126 (16) | 135 (17) |
| Hypertension treatment | 428 (39.2) | 523 (57.3) | 133 (36.8) | 1369 (65.6) | 139 (68.8) |
| Stage I Hypertension | 443 (40.6) | 538 (58.9) | 188 (52.2) | 1478 (70.8) | 160 (79.2) |
| Prevalent diabetes | 74 (6.8) | 118 (12.9) | 46 (12.8) | 272 (13.0) | 24 (11.9) |
| Prevalent CVD | 114 (10.4) | 106 (11.6) | 41 (11.3) | 852 (40.9) | 36 (17.8) |
| Current smoker | 97 (8.9) | 53 (5.8) | 39 (10.7) | 40 (1.9) | 4 (2.0) |
| Body mass index, Median (Q1, Q3), kg/m<sup>2</sup> | 28.3 (25.3, 31.8) | 27.3 (24.7, 29.9) | 27.3 (24.7, 30.7) | 26.8 (24.7, 29.4) | 27.3 (24.6, 31.2) |
| Sleeping pill use | 255 (23.4) | 214 (23.6) | 50 (14.2) | 421 (20.2) | 46 (22.8) |
| Antidepressant use | 67 (6.1) | 48 (5.3) | 10 (2.8) | 155 (7.4) | 11 (5.5) |
| Sedative use | 58 (5.3) | 72 (7.9) | 13 (3.6) | 140 (6.7) | 31 (15.4) |
| APOE e4 carrier | 309 (28.3) | 218 (23.9) | 81 (22.3) | 484 (23.2) | 22 (10.9)** |
| Incident all cause Dementia | 289 (26.5%) | 538 (58.9%) | 73 (20.1%) | 75 (3.6) | 23 (11.4) |
| Median follow-up time, years (Q1, Q3) | 19.2 (13.8, 21.6) | 8.5 (4.1, 13.6) | 15.7 (9.8, 21.1) | 10.5 (7.6, 11.4) | 4.8 (4.1, 5.2) |

\*Values are n (%) unless otherwise specified as mean (SD) or median [Q1, Q3] for non-normally distributed variables. \*\* *APOE e4* carrier status was calculated as the *percentage among those for whom their status* was known in SOF. ARIC, Atherosclerosis Risk in Communities study; CHS, Cardiovascular Health Study; FHS, Framingham Heart Study; MrOS, Osteoporotic Fractures in Men Study; SOF, Study of Osteoporotic Fractures; CVD, cardiovascular disease.

### Meta-analysis

Study-level estimates were pooled centrally in random effects meta-analyses. The Sidik-Jonkman estimator method was used to calculate the heterogeneity variance τ^2^ and the classic method was used to calculate the 95% CIs around the pooled effect. The Higgins I^2^ test was implemented to test for heterogeneity in effect sizes.^19^ Statistical tests were all two-sided. All results were considered significant if p<0.05.

### Exploration of effect modification

Since sex^20, 21^ and *APOE*^22^ allele carrier status are associated with both sleep and dementia risk, we explored effect modification by *APOE e4* allele carrier status (non-*e4* carrier vs. at least one copy of *e4*) and sex (men vs. women) by including interaction terms in age and sex adjusted models. In the presence of a significant interaction (p<0.05), results were stratified at each level of the moderating variable. Interaction results were not pooled in meta-analysis. Rather, we interpreted patterns that were evident across studies. Sex was not examined as a moderating variable in MrOS or SOF since these cohorts were exclusively men and women, respectively, and *APOE* genotype was not examined as a moderating variable in SOF (since it was not available on all participants).

### Secondary analysis

We performed a secondary random effects meta-analysis restricted to ARIC, CHS, and the FHS based on these three cohorts having PSGs performed at the same time (from 1995-1998) in a methodologically consistent manner as part of the multicenter Sleep Heart Health Study (SHHS)^16^. Moreover, these cohorts had the most similar methods in terms of dementia surveillance and adjudication.

### Sensitivity analysis

To account for the potential confounding influence of OSA on sleep macro-architecture measures, the primary analyses were repeated including adjustments with the addition of the AHI.

## RESULTS

Sample demographics and sleep architecture measures across cohorts are presented in Table 1 and 2, respectively. Overall, 4,657 participants were included in the analysis (30% were women [owing to the large MrOS study being male only]; mean age (weighted by cohort size) was 74 ± 12 years and ranged from 65-83 years at the time of PSG; 3.9% were Black; 95.6% were White, and 0.5% were categorized as other race or ethnicity. Across cohorts, 10.8% of participants did not have a high school degree and 21.2% of participants reported using sleeping pills regularly. In total, there were 998 (21.4%) incident dementia cases across cohorts, with the highest percentage of cases reported in the CHS (58.9%), one of the oldest cohorts with long follow-up. The lowest percentage of dementia cases was in the MrOS cohort (3.6%), which included only men. The median follow-up time ranged between 4.8 to 19.2 years across cohorts. Sleep characteristics for each cohort are presented in Table 2. Sleep stages were mostly similar between cohorts, with the exception of MrOS which had the highest levels of N1 and N2% and lowest levels of N3%. SME was similar between ARIC, CHS and FHS (ranging between 85-87%), but lowest in MrOS and SOF (range 78-82%). Similarly, WASO ranged between 54-63 minutes across ARIC, CHS and FHS and was highest in MrOS and SOF (76 to 101 minutes). Across cohorts, the average AHI ranged between 6-8 events/hour. The proportion who experienced short sleep duration of ≤6 hours per night ranged from 43-52%.

**Table 2:** Sleep Characteristics.

| **Median (Q1, Q3)\*** | **ARIC**<br>**N=1,092** | **CHS**<br>**N=913** | **FHS**<br>**N=363** | **MrOS**<br>**N=2,087** | **SOF**<br>**N=202** |
| --- | --- | --- | --- | --- | --- |
| N1 (%) | 5.1 (3.1, 7.7) | 4.7 (2.7, 7.3) | 4.8 (3.0, 7.3) | 6.0 (4.0, 8.0) | 4.1 (2.7, 5.9) |
| N2 (%) | 56.0 (11.5) | 57.9 (12.6) | 56.75 (12.0) | 62.6 (9.5) | 55.0 (12.5) |
| N3 (%) | 17.5 (8.3, 25.4) | 16.3 (7.3, 25.8) | 17.7 (8.8, 26.3) | 10.3 (4.1, 17.0) | 19.6 (12.6, 29.1) |
| REM (%) | 20.4 (5.9) | 18.7 (6.3) | 19.5 (5.9) | 19.3 (6.5) | 18.6 (6.8) |
| Sleep Maintenance Efficiency (%) | 87.4 (80.6, 91.6) | 85.0 (76.7, 90.8) | 87.0 (80.0, 92.1) | 78.5 (70.4, 85.0) | 82.2 (73.4, 88.7) |
| Wake After Sleep Onset (min) | 54.0 (34.5, 85.0) | 63.5 (37.5, 101.3) | 55.5 (32.5, 87.5) | 101.0 (66.0, 145.0) | 76.0 (47.5, 116.5) |
| Apnea hypopnea index (no. events/hour) | 6.2 (2.3, 13.9) | 7.3 (2.8, 15.4) | 6.5 (2.2, 14.3) | 8.2 (3.2, 17.1) | 6.4 (2.5, 13.2) |
| Total sleep time ≤6 hours, n (%) | 488 (44.7) | 471 (51.6) | 157 (43.3) | 1038 (49.7) | 104 (51.4) |

\*Values are median (Q1, Q3) for non-normally distributed data or mean (SD) for normally distributed data, unless specified as n (%). N1, stage 1 non-rapid eye movement sleep; N2, stage 2 non-rapid eye movement sleep; N3, stage 3 non-rapid eye movement sleep; REM, rapid eye movement sleep.

### Sleep and dementia risk

Figure 1 presents the meta-analysed results, summarizing the association between sleep exposures and incident all-cause dementia. In the majority of cohorts, higher N3% (Fig 1c) and lower REM% (Fig 1d) trended toward increased dementia risk. However, pooled estimates revealed no statistically significant associations between these sleep exposures and dementia risk. Higher SME% (Fig 1e) and lower WASO (Fig 1f) trended towards reduced dementia risk in some cohorts (e.g., ARIC, FHS and SOF); however, the overall pooled association with dementia risk was not statistically significant. Additionally, heterogeneity in effect estimates between studies was moderate to substantial in these models (SME% I^2^ = 50% [95% CI 0.0%, 82%]; WASO I^2^ = 72% [95% CI 29%, 89%]).

**Figure 1:**
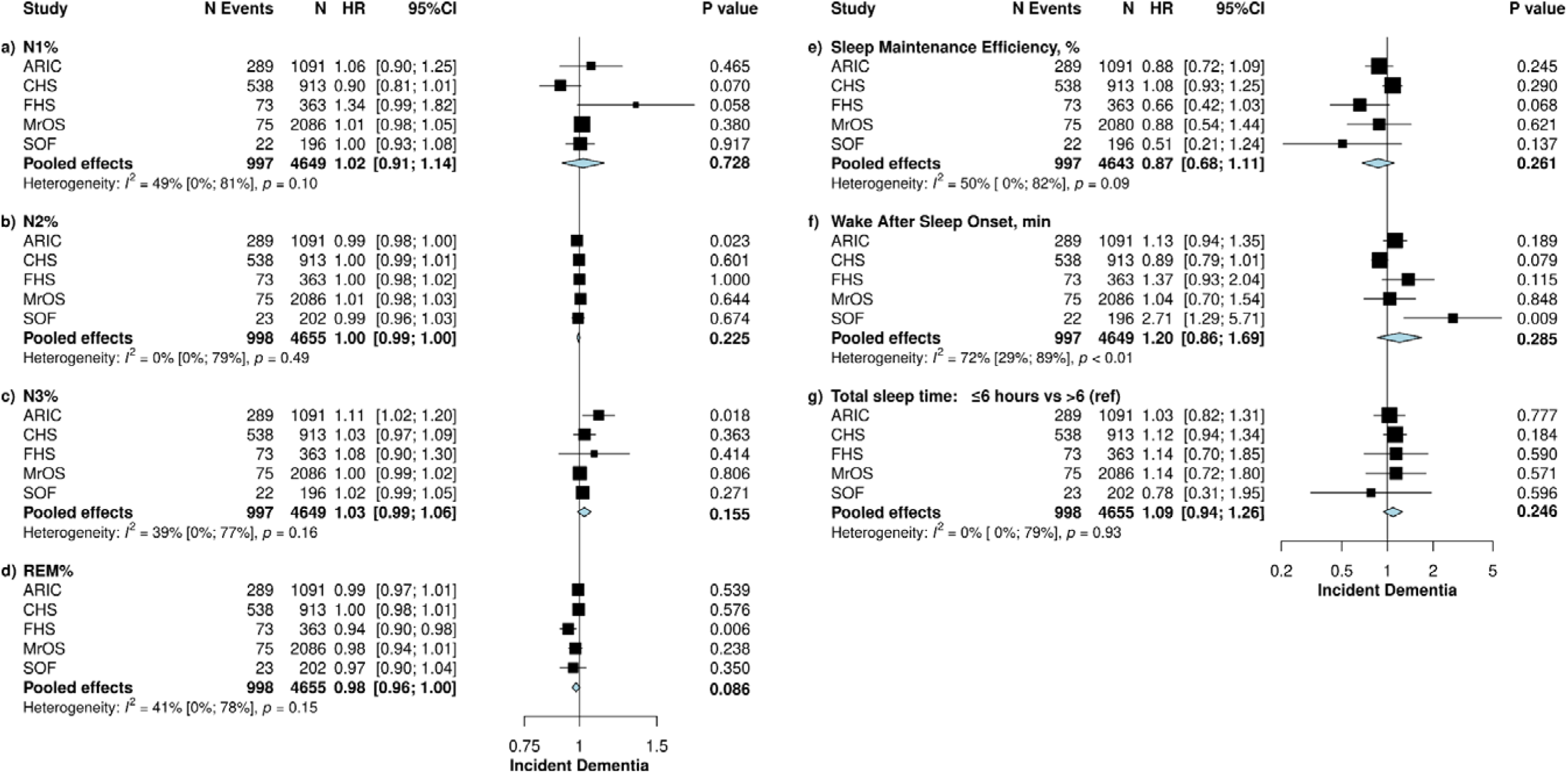
Pooled association between sleep macro-architecture measures and incident dementia. Figure depicts the meta-analysis with forest plot. All results were adjusted for age (years), sex (men vs. women), BMI (kg/m^2^), anti-depressant use (yes vs no), sedative use (yes vs no), and *APOE e4* status (non e4 carrier vs. at least one copy of e4). Cohort studies included: ARIC, Atherosclerosis Risk in Communities study; CHS, Cardiovascular Health Study; FHS, Framingham Heart Study; MrOS, Osteoporotic Fractures in Men Study; SOF, Study of Osteoporotic Fractures. The sleep exposures in each model included: N1, non-rapid eye movement sleep stage 1; N2, non-rapid eye movement sleep stage 2; N3, non-rapid eye movement sleep stage 3; REM, rapid eye movement sleep; WASO, Wake after sleep onset; SME, sleep maintenance efficiency. Note that, square root transformation was applied to N1% and N3% and natural log transformation was applied to SME%, WASO and the AHI due to skewed distributions of these sleep metrics. Dementia case numbers are presented for each cohort with hazard ratio (HR) and 95% confidence intervals (95% CI) for dementia risk. Heterogeneity in effect sizes was determined via the Higgins I^2^ test. Statistical significance, p<0.05.

In the secondary analysis limiting to CHS, FHS and ARIC from the SHHS, there were 2,367 participants and 900 incident dementia cases. Compared to the primary analyses, the age range was slightly younger (range 65 to 77 years; weighted mean age 70.5 years), a higher proportion were women (51.6%), and the average follow-up time was longer (range 8.5 to 19.2 years). In these analyses restricting to three cohorts, higher N3% was marginally associated with increased dementia risk (HR, 1.06; 95% CI, 1.00 - 1.12, p<0.05), such that for every percentage increase in N3 sleep there was a 6% increase in dementia risk (Figure 2). There were no other associations identified with the other sleep exposure variables.

**Figure 2:**
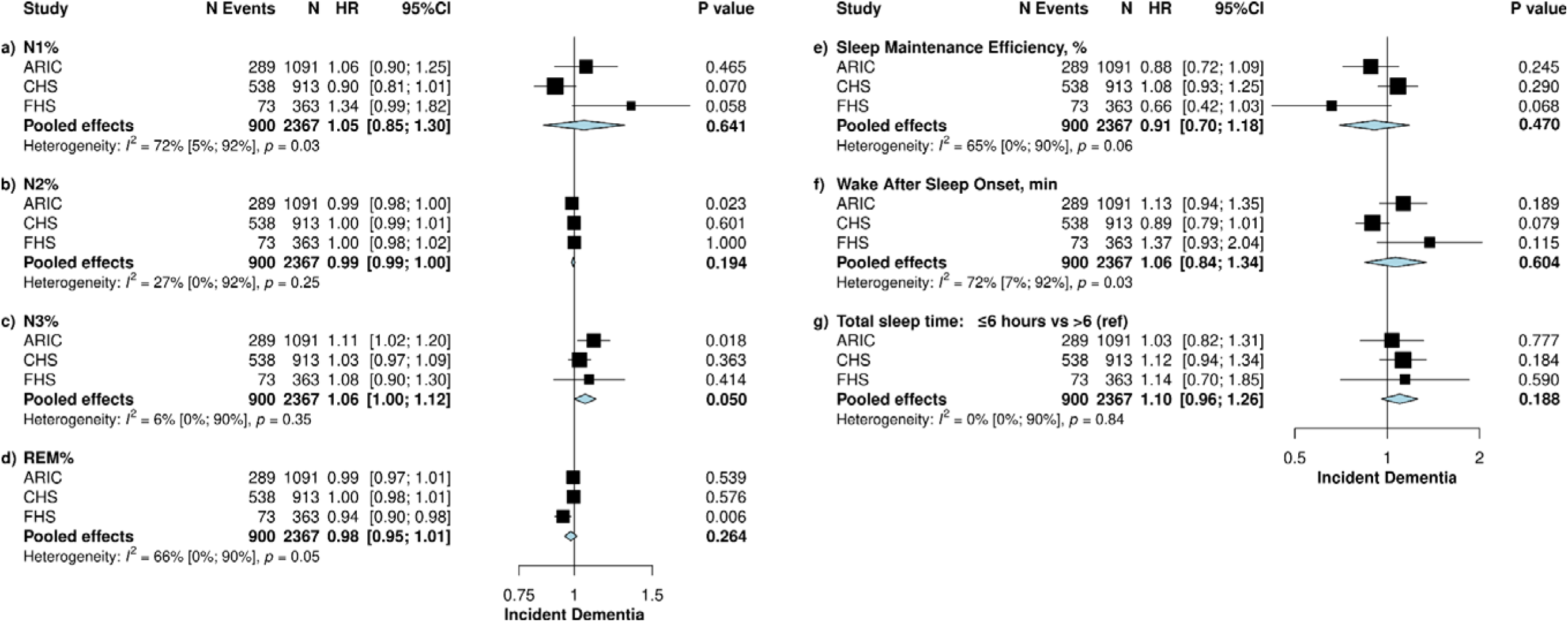
Pooled association between sleep macro-architecture measures and incident dementia – secondary analysis restricted to ARIC, CHS, FHS cohorts. Figure depicts the meta-analysis with forest plot limited to three cohorts. All results were adjusted for age (years), sex (men vs. women), BMI (kg/m^2^), anti-depressant use (yes vs. no), sedative use (yes vs. no), and *APOE e4* status (non e4 carrier vs. at least one copy of e4). Cohort studies included: ARIC, Atherosclerosis Risk in Communities study; CHS, Cardiovascular Health Study and the FHS, Framingham Heart Study. The sleep exposures in each model included: N1, non-rapid eye movement sleep stage 1; N2, non-rapid eye movement sleep stage 2; N3, non-rapid eye movement sleep stage 3; REM, rapid eye movement sleep; WASO, Wake after sleep onset; SME, sleep maintenance efficiency. Note that, square root transformation was applied to N1% and N3% and natural log transformation was applied to SME%, WASO and the AHI due to skewed distributions of these sleep metrics. Dementia case numbers are presented for each cohort with hazard ratio (HR) and 95% confidence intervals (95% CI) for dementia risk. Heterogeneity in effect sizes was determined via the Higgins I^2^ test. Statistical significance, p<0.05.

### Moderation analysis

Results for the moderation analysis by *APOE e4* status and sex are presented in Supplementary Tables 2-5. There was no consistent pattern of moderation of *APOE e4* status or sex between any of the sleep measures and dementia incidence across cohorts.

### Sensitivity analysis

Additional adjustment for the AHI did not meaningfully alter the results (Supplementary Table 6).

## DISCUSSION

To our knowledge, this is the largest initiative to investigate the association between PSG-derived sleep macro-architecture measures and risk of dementia. Across five US cohorts, the overall pooled effects showed no association between sleep variables and incident dementia, although some sleep stage percentages and measures of sleep disruption trended toward increased dementia risk. Further, the effect modification analysis showed few and inconsistent interactions of sex and *APOE e4* positivity on associations between sleep and dementia. There was also moderate to large heterogeneity between study effects for some sleep metrics. Of note, when data were restricted to the three SHHS cohorts, there was an unexpected suggestion that higher N3% was associated with greater increased dementia risk but not for other stages.

N3 sleep is important for memory consolidation^23, 24^ and animal studies indicate that slow wave activity (occurring at the delta frequency 0.5-4 Hz), which is dominant during N3 sleep, plays an important role in the glymphatic clearance of amyloid-β^7^. Thus, we hypothesized that lower N3% may be one of the strongest predictors of dementia risk. In contrast to our expectations, our primary results suggest that individual difference in N3 quantity at a discrete time point may not be meaningful for dementia prediction. In line with these findings, the initial analysis of the SDC cohorts also showed no association between sleep stage durations and cognition^11^.

There are several explanations for our findings. Importantly, the measure of N3% does not completely capture the full range and detail of EEG oscillatory activity (e.g., including Up and Down states of increased and quiet activity) reflective of variations in cortical rhythms believed to be important for synaptic plasticity and memory consolidation^25^. That is, within scored N3 sleep, the duration of slow waves, their coupling with spindles, and frequency power vary. Further, within the delta frequency band, slow frequencies (0.5-1 Hz) vs higher frequencies (up to 4 Hz) have inverse associations with cognitive outcomes^13^. Previous studies that utilized spectral analysis have shown that the duration of slow wave oscillations and the ratio of slow (<1 Hz) to delta waves (0.5 to 4 Hz), not N3 duration, were associated with cognitive performance^13^, which suggests the need to characterize multiple aspects of slow wave activity, as well as EEG features such as spindles and K complexes.

Another explanation for our findings could be the age range at which participants were studied. As N3 sleep declines with age, individual differences at younger ages than studied here may be more relevant to the study of late-life dementia risk (e.g., a lower percentage of N3 sleep at an earlier age may be indicative of accelerated brain aging). It is also possible that a loss or gain in N3% over time is more important for determining dementia risk, rather than differences between individuals at a given time point. Recent data from the FHS showed that a 1% decline in N3% per year was associated with a 27% increased risk of dementia^22^.

The finding that higher N3% was marginally associated with increased dementia risk in the analysis restricted to the three original SHHS cohorts should not be overinterpreted, given that it was not significant in the primary analyses and the large number of comparisons conducted. Nevertheless, there are a number of mechanisms that could explain this association. N3 sleep is a highly reactive sleep stage. Higher N3 sleep may reflect high homeostatic sleep pressure due to prior night sleep deprivation. That is, higher N3% captured by the single overnight sleep study may serve as a proxy for irregular sleep patterning or chronic sleep deprivation. Alternatively, our findings may reflect the capacity of central mechanisms to make a compensatory increase in the proportion of N3 sleep to protect the brain. It has been proposed that N3 sleep may be responsive to wake-dependent build-up of metabolic and oxidative by-products, whereby slow wave activity increases to counterbalance amyloid-β aggregation^26^. Those with higher amounts of N3 sleep may reflect individuals with elevated levels of amyloid. Speculatively, in select people, loss of the ability for N3 sleep to accommodate the accumulation of amyloid could reflect the tipping point at which individuals become vulnerable to neurodegeneration and subsequent dementia. However, further longitudinal studies with sleep assessed at multiple time points are required to confirm this contention.

In contrast to previous studies showing that reduced REM% is associated with cognitive decline^27^ and dementia risk^10^, this study found no association between REM% and incident dementia. The possible null findings in this meta-analysis may be due to the older age range studied in the overall sample. Unlike N3 sleep, REM sleep sees little decline with older age^1^, resulting in a smaller range of REM changes which may be too small to detect a difference.

Our previous results in the SDC cohort^11^, as well as findings from other studies utilising actigraphy^28^, indicate that higher levels of sleep disruption (e.g., lower sleep efficiency and high WASO) are associated with poorer cognition. Although there was a nominal tendency for sleep disruption measures to associate with dementia risk in some cohorts, the pooled effects in this study showed no significant associations. We did observe that higher amounts of WASO were associated with significantly increased dementia risk in the SOF cohort, which was the oldest cohort with the shortest duration of follow-up. Thus, it is possible that reverse causation may underlie this relationship. Measures of sleep disruption at multiple time points across the lifespan would be helpful to further determine the temporal association between WASO and dementia risk.

We also investigated whether any of the study effects were influenced by *APOE* or sex. While *APOE e4* carriage is the most important genetic risk factor for dementia^29^, and dementia risk is highest among women ^30^, evidence to show that these factors (*APOE e4* carriage or sex) moderated any of the sleep-dementia associations was inconsistent between studies. However, these interactions could be dependent on cohort-specific characteristics (e.g., age or sex distributions). Thus, investigating these moderation effects may require a larger sample size (with a higher number of dementia cases) to truly tease out these relationships if they exist.

This study has a number of strengths including the large sample size, long prospective follow-up duration, representation from five different cohorts and use of methodologically consistent PSG recordings. However, there were several limitations. Firstly, not all studies adjudicated dementia using DSM criteria. To address this in some manner, we performed a sensitivity analysis that contained only the original SHHS cohorts, which had similar dementia adjudication methods. In this analysis, we saw HRs of mostly similar in magnitude and direction for all sleep measures. Secondly, sleep was measured over only a single night, which may not fully represent habitual sleep patterns and sleep architecture. Thus, future studies that utilize PSG recordings across several nights may provide a more robust characterization of sleep architecture. Also, we may not have assessed sleep at the optimal life stage to explore associations with dementia.

### Implications and Conclusions

Sleep has been implicated in many mechanisms that are related to dementia, such as memory formation^31^, glymphatic clearance^7^, and vascular brain health^32^. However, the role of poor sleep as a dementia risk factor remains poorly understood. The current findings suggest that sleep stage quantity or basic sleep disruption measures obtained at a single time point may not be useful for predicting long-term dementia risk overall. In light of this, research efforts could be directed to explore more precise neurophysiological measures of sleep, including quantitative measures of oscillations and dynamic changes of the EEG across the sleep period and the distributions of sleep spindles. Moreover, rather than using individual metrics as exposures, more advanced statistical techniques could be used to derive different sleep phenotypes from combinations of individual metrics. Also, when considering other recent findings^22^, understanding the trajectories of sleep with aging, rather than single point measurements, may be more informative for dementia risk. This study also highlights that there is marked variability in sleep-dementia associations between different cohorts. Identification of the drivers of this heterogeneity will be an important next step, as there may be certain factors that protect against dementia in the face of poor sleep. In other words, individual sleep metrics may be more predictive in specific subpopulations.

In conclusion, sleep macro-architecture measures did not associate with dementia risk. Although N3 sleep plays a role in glymphatic clearance^7^ and declining N3 sleep has been linked with dementia^22^, individual differences in N3 sleep do not have a straightforward association with dementia risk. Future studies examining micro-architectural measures of sleep, how sleep metrics change over time, and N3 associations with amyloid burden will be important for further elucidating sleep and dementia relationships.

## ACKNOWLEDGEMENTS

**SDC:** The SDC is funded by the NIA (R01 AG062531). This work is also made possible by grants from the NIA to the Cross Cohorts Consortium (CCC) (AG059421) and by the Cohorts for Age and Aging Research in Genomic Epidemiology (CHARGE) infrastructure grant from the NHLBI (HL105756).

**FHS:** This work was made possible by grants from the National Institutes of Health (N01-HC-25195, HHSN268201500001I, 75N92019D00031) and the National Institute on Aging (AG059421, AG054076, AG049607, AG033090, AG066524, NS017950, P30AG066546, UF1NS125513).

**ARIC:** The ARIC portion of the SHHS was supported by National Heart, Lung, and Blood Institute cooperative agreements U01HL53934 (University of Minnesota) and U01HL64360 (Johns Hopkins University). The Atherosclerosis Risk in Communities study has been funded in whole or in part with Federal funds from the National Heart, Lung, and Blood Institute, National Institutes of Health, Department of Health and Human Services, under Contract nos. (75N92022D00001, 75N92022D00002, 75N92022D00003, 75N92022D00004, 75N92022D00005). The authors thank the staff and participants of the ARIC study for their important contributions.

ARIC Neurocognitive (ARIC-NCS) for selected components of Visit 5 and 7 and all of Visits 6-8 The Atherosclerosis Risk in Communities Study is carried out as a collaborative study supported by National Heart, Lung, and Blood Institute contracts (75N92022D00001, 75N92022D00002, 75N92022D00003, 75N92022D00004, 75N92022D00005). The ARIC Neurocognitive Study is supported by U01HL096812, U01HL096814, U01HL096899, U01HL096902, and U01HL096917 from the NIH (NHLBI, NINDS, NIA and NIDCD). The authors thank the staff and participants of the ARIC study for their important contributions.

**CHS:** This research was supported by contracts HHSN268201200036C, HHSN268200800007C, HHSN268201800001C, N01HC55222, N01HC85079, N01HC85080, N01HC85081, N01HC85082, N01HC85083, N01HC85086, 75N92021D00006, and grants U01HL080295 and U01HL130114 from the National Heart, Lung, and Blood Institute (NHLBI), with additional contribution from the National Institute of Neurological Disorders and Stroke (NINDS). Support for cognitive measures was provided by R01AG15928. Additional support was provided by R01AG023629 from the National Institute on Aging (NIA). A full list of principal CHS investigators and institutions can be found at CHS-NHLBI.org.

**MrOS:** MrOS is supported by National Institutes of Health funding. The following institutes provide support: the National Institute on Aging (NIA), the National Institute of Arthritis and Musculoskeletal and Skin Diseases (NIAMS), the National Center for Advancing Translational Sciences (NCATS), and NIH Roadmap for Medical Research under the following grant numbers: U01 AG027810, U01 AG042124, U01 AG042139, U01 AG042140, U01 AG042143, U01 AG042145, U01 AG042168, U01 AR066160, and UL1 TR0 The National Heart, Lung, and Blood Institute (NHLBI) provides funding for the MrOS Sleep ancillary study “Outcomes of Sleep Disorders in Older Men” under the following grant numbers: R01 HL071194, R01 HL070848, R01 HL070847, R01 HL070842, R01 HL070841, R01 HL070837, R01 HL070838, and R01 HL07083900128.

**SOF:** SOF is supported by National Institutes of Health funding. The National Institute on Aging (NIA) provides support under the following grant numbers: R01 AG005407, R01 AR35582, R01 AR35583, R01 AR35584, R01 AG005394, R01 AG027574, R01 AG027576, and R01 AG026720.

**Other:** Dr Yiallourou are funded by the American Alzheimer’s Association (AARG-NTF-22-971405). Dr. Pase is supported by the National Health and Medical Research Council of Australia (GTN2009264; GTN1158384). Dr. Cavuoto and Dr. Pase are supported by a Dementia Australia Research Foundation award (Lucas’ Papaw Remedies Project Grant). Dr. Baril is funded by grants from the Alzheimer Society of Canada, Canadian Institutes of Health Research, and Sleep Research Society Foundation. Drs. Seshadri and Himali are partially supported by the South Texas Alzheimer’s Disease Center (1P30AG066546-01A1) and The Bill and Rebecca Reed Endowment for Precision Therapies and Palliative Care. Dr. Seshadri is also supported by an endowment from the Barker Foundation as the Robert R Barker Distinguished University Professor of Neurology, Psychiatry and Cellular and Integrative Physiology and Dr. Himali by an endowment from the William Castella family as William Castella Distinguished University Chair for Alzheimer’s Disease Research. Dr. Redline is partially funded by NIA AG 070867. Dr. Yaffe is partially funded by NIA R35 AG071916.

Dr. Lutsey is partially supported by K24 HL159246. Dr. Gottesman is supported by the NINDS Intramural Research Program. Dr. Purcell is partially funded by NIH NHLBI R01HL146339, NIH NIA 070867, and NIH NIMHD MD012738.

We thank the participants for dedicating their time to our research. We also thank the researchers involved in data collection and obtaining funding, including Anne Newman (MD, MPH) and John Robbins (MD, MHS) from the Cardiovascular Health Study, who received no compensation for the current article.

## DATA AVAILABILITY

The SDC cohorts makes de-identified phenotypic and genetic data available through the online repositories BioLINCC and dbGaP, respectively. Sleep data are available via the National Sleep Research Resource (NSRR) https://sleepdata.org/

## REFERENCES

1. Edwards BA, O’Driscoll DM, Ali A, Jordan AS, Trinder J, Malhotra A. Aging and sleep: physiology and pathophysiology. Semin Respir Crit Care Med. Oct 2010;31(5):618–33. doi:10.1055/s-0030-1265902

2. Tononi G, Cirelli C. Sleep and synaptic homeostasis: a hypothesis. Brain Res Bull. Dec 15 2003;62(2):143–50. doi:10.1016/j.brainresbull.2003.09.004

3. Wunderlin M, Zust MA, Feher KD, Kloppel S, Nissen C. The role of slow wave sleep in the development of dementia and its potential for preventative interventions. Psychiatry Res Neuroimaging. Dec 30 2020;306:111178. doi:10.1016/j.pscychresns.2020.111178

4. Lucey BP, McCullough A, Landsness EC, et al. Reduced non-rapid eye movement sleep is associated with tau pathology in early Alzheimer’s disease. Sci Transl Med. Jan 9 2019;11(474)doi:10.1126/scitranslmed.aau6550

5. Winer JR, Mander BA, Helfrich RF, et al. Sleep as a Potential Biomarker of Tau and beta-Amyloid Burden in the Human Brain. J Neurosci. Aug 7 2019;39(32):6315–6324. doi:10.1523/JNEUROSCI.0503-19.2019

6. Miao A, Luo T, Hsieh B, et al. Brain clearance is reduced during sleep and anesthesia. Nat Neurosci. Jun 2024;27(6):1046–1050. doi:10.1038/s41593-024-01638-y

7. Xie L, Kang H, Xu Q, et al. Sleep drives metabolite clearance from the adult brain. Science. Oct 18 2013;342(6156):373-7. doi:10.1126/science.1241224

8. Sprecher KE, Koscik RL, Carlsson CM, et al. Poor sleep is associated with CSF biomarkers of amyloid pathology in cognitively normal adults. Neurology. Aug 1 2017;89(5):445–453. doi:10.1212/WNL.0000000000004171

9. Yaffe K, Laffan AM, Harrison SL, et al. Sleep-disordered breathing, hypoxia, and risk of mild cognitive impairment and dementia in older women. JAMA. Aug 10 2011;306(6):613–9. doi:10.1001/jama.2011.1115

10. Pase MP, Himali JJ, Grima NA, et al. Sleep architecture and the risk of incident dementia in the community. Neurology. Sep 19 2017;89(12):1244–1250. doi:10.1212/WNL.0000000000004373

11. Pase MP, Harrison S, Misialek JR, et al. Sleep Architecture, Obstructive Sleep Apnea, and Cognitive Function in Adults. JAMA Netw Open. Jul 3 2023;6(7):e2325152. doi:10.1001/jamanetworkopen.2023.25152

12. Cavuoto MG, Ong B, Pike KE, Nicholas CL, Bei B, Kinsella GJ. Objective but not subjective sleep predicts memory in community-dwelling older adults. J Sleep Res. Aug 2016;25(4):475–85. doi:10.1111/jsr.12391

13. Djonlagic I, Mariani S, Fitzpatrick AL, et al. Macro and micro sleep architecture and cognitive performance in older adults. Nat Hum Behav. Jan 2021;5(1):123–145. doi:10.1038/s41562-020-00964-y

14. Swanson LM, Hood MM, Hall MH, et al. Associations between sleep and cognitive performance in a racially/ethnically diverse cohort: the Study of Women’s Health Across the Nation. Sleep. Feb 12 2021;44(2)doi:10.1093/sleep/zsaa182

15. von Elm E, Altman DG, Egger M, et al. Strengthening the Reporting of Observational Studies in Epidemiology (STROBE) statement: guidelines for reporting observational studies. BMJ. Oct 20 2007;335(7624):806–8. doi:10.1136/bmj.39335.541782.AD

16. Quan SF, Howard BV, Iber C, et al. The Sleep Heart Health Study: design, rationale, and methods. Sleep. Dec 1997;20(12):1077–85.

17. Whitney CW, Gottlieb DJ, Redline S, et al. Reliability of scoring respiratory disturbance indices and sleep staging. Sleep. Nov 1 1998;21(7):749–57. doi:10.1093/sleep/21.7.749

18. Redline S, Sanders MH, Lind BK, et al. Methods for obtaining and analyzing unattended polysomnography data for a multicenter study. Sleep Heart Health Research Group. Sleep. Nov 1 1998;21(7):759–67.

19. Higgins JP, Thompson SG. Quantifying heterogeneity in a meta-analysis. Stat Med. Jun 15 2002;21(11):1539–58. doi:10.1002/sim.1186

20. Gong J, Harris K, Lipnicki DM, et al. Sex differences in dementia risk and risk factors: Individual-participant data analysis using 21 cohorts across six continents from the COSMIC consortium. Alzheimers Dement. Aug 2023;19(8):3365–3378. doi:10.1002/alz.12962

21. Redline S, Kirchner HL, Quan SF, Gottlieb DJ, Kapur V, Newman A. The effects of age, sex, ethnicity, and sleep-disordered breathing on sleep architecture. Arch Intern Med. Feb 23 2004;164(4):406–18. doi:10.1001/archinte.164.4.406

22. Himali JJ, Baril AA, Cavuoto MG, et al. Association Between Slow-Wave Sleep Loss and Incident Dementia. JAMA Neurol. Oct 30 2023;doi:10.1001/jamaneurol.2023.3889

23. Diekelmann S, Born J. The memory function of sleep. Nat Rev Neurosci. Feb 2010;11(2):114–26. doi:10.1038/nrn2762

24. Tononi G, Cirelli C. Sleep function and synaptic homeostasis. Sleep Med Rev. Feb 2006;10(1):49-62. doi:10.1016/j.smrv.2005.05.002

25. Torao-Angosto M, Manasanch A, Mattia M, Sanchez-Vives MV. Up and Down States During Slow Oscillations in Slow-Wave Sleep and Different Levels of Anesthesia. Front Syst Neurosci. 2021;15:609645. doi:10.3389/fnsys.2021.609645

26. Mander BA, Winer JR, Jagust WJ, Walker MP. Sleep: A Novel Mechanistic Pathway, Biomarker, and Treatment Target in the Pathology of Alzheimer’s Disease? Trends Neurosci. Aug 2016;39(8):552–566. doi:10.1016/j.tins.2016.05.002

27. Song Y, Blackwell T, Yaffe K, et al. Relationships between sleep stages and changes in cognitive function in older men: the MrOS Sleep Study. Sleep. Mar 1 2015;38(3):411–21. doi:10.5665/sleep.4500

28. Yaffe K, Falvey CM, Hoang T. Connections between sleep and cognition in older adults. Lancet Neurol. Oct 2014;13(10):1017–28. doi:10.1016/S1474-4422(14)70172-3

29. Liu CC, Liu CC, Kanekiyo T, Xu H, Bu G. Apolipoprotein E and Alzheimer disease: risk, mechanisms and therapy. Nat Rev Neurol. Feb 2013;9(2):106–18. doi:10.1038/nrneurol.2012.263

30. G. B. D. 2019 Dementia Forecasting Collaborators. Estimation of the global prevalence of dementia in 2019 and forecasted prevalence in 2050: an analysis for the Global Burden of Disease Study 2019. Lancet Public Health. Feb 2022;7(2):e105-e125. doi:10.1016/S2468-2667(21)00249-8

31. Rasch B, Born J. About sleep’s role in memory. Physiological reviews. 2013;

32. Baril A-A, Beiser AS, Mysliwiec V, et al. Slow-wave sleep and MRI markers of brain aging in a community-based sample. Neurology. 2021;96(10):e1462–e1469.

