## Supplementary material for "Sleep macro-architecture and dementia risk in adults: Meta-analysis of 5 cohorts from the Sleep and Dementia Consortium"

**Supplementary Methods:**

**SDC Cohort description surveillance for Dementia Case Ascertainment**

**i. ARIC:**

The Atherosclerotic Risk in Communities (ARIC) study is a large, prospective study established in 1987 to study cardiovascular disease in four geographically and racially diverse US communities. Participants recruited to the Sleep Heart Health Study were restricted to two communities in ARIC where the majority self-reported as white. Participants underwent dementia surveillance from the time of their baseline sleep assessment (1996-1998) through to 2019, with a median follow-up time of 19.2 (Q1: 13.8, Q3: 21.6) years. Surveillance for dementia case ascertainment in ARIC has been described in detail previously,^1^ and includes several approaches. First, participants underwent a detailed neurocognitive assessment at visit 5 (2011-2013) and all subsequent visits (visit 6: 2016-2017; visit 7: 2018-2019). At visit 5, a selected subset also received a neurological exam and brain magnetic resonance imaging (MRI). Second, two validated phone-based cognitive assessment tools have been used. A modified telephone interview for cognitive status (TICSm) was conducted among participants who at the time of visit 5 were alive but unable or unwilling to participate in an in-person exam. Informants were queried when appropriate. From visit 5 forward, participants were called twice yearly and screened for dementia using the Six Item Screener accompanied by administration of the AD8 dementia screener to informants when appropriate. Lastly, the full cohort has had continuous surveillance for hospitalization and death since 1987. Dementia hospitalization and death diagnosis codes were also used to ascertain dementia. The ARIC Neurocognitive Classification Committee reviewed each potential dementia case. Dementia diagnosis was based on Diagnostic and Statistical Manual of Mental Disorders, 5th Edition (DSM-5)^2^.

**ii. CHS:**

The Cardiovascular Heart Study (CHS) is a prospective population-based cohort study of cardiovascular disease in adults aged over 65 years in four US field centers. For participants who participated in the CHS Cognition Study, CHS ascertained dementia through June 30, 1999 using a standardized protocol. The median follow-up time for dementia surveillance was 8.5 (Q1: 4.1, Q3: 13.6) years. All probable dementia cases were formally reviewed by an adjudication committee composed of expert neurologists and psychiatrists; details of this protocol have been previously published^3^. DSM-IV criteria^4^ was used for the clinical diagnosis of dementia and was based on a progressive or static cognitive deficit of sufficient severity to affect the subjects’ activities of daily living and history of normal intellectual function before the onset of cognitive abnormalities. For CHS Cognition Study participants at the Pittsburgh site, this dementia ascertainment protocol was continued through June 3, 2013. For all other participants (and CHS Cognition Study participants who were without dementia at the time monitoring ended), multiple sources of data were used to identify incident dementia through June 2015. Data sources included medication use (e.g., donepezil, galantamine, memantine, rivastigmine, tacrine), International Classification of Diseases-9 codes (290.xx, 294.xx, 331.0, 331.1, 331.2, 331.82, 331.83, 331.9, and 438.0), needing a proxy due to cognitive problems, and adjudicated cause of death listed as dementia on death certificates. Participants were also required to have impairment in two cognitive domains, which did not necessarily include memory.

**iii. FHS:**

The Framingham Heart Study (FHS) is an ongoing, longitudinal, community-based cohort study initiated in 1948 to investigate risk factors for cardiovascular disease. This study uses data from the Offspring cohort (followed since 1971) and a multi-ethnic cohort of racially diverse adults was recruited in 1994-98, tested in parallel with the Offspring cohort. FHS participants are under continuous surveillance for incident dementia, the methods for which are detailed elsewhere^5^. Surveillance for this study was from the time of the baseline sleep study (1995-1998) through to 2020, with a median follow-up time of 15.6 (Q1: 9.7, Q3: 21.0) years. In brief, cognitive screening is performed at each FHS examination cycle using the Mini-Mental State Examination (MMSE) augmented by extensive neuropsychological testing at selected examinations. The MMSE is used to flag suspected cognitive impairment if (1) performance falls below education-based cutoff scores, (2) a decline of 3 or more points is observed between consecutive examinations, or (3) a decrease of 5 or more points is observed from the participant's highest past MMSE score. Participants are also flagged for suspected cognitive impairment following referrals from FHS investigators or outside practitioners if concern is expressed by the participant or the family. Once flagged for suspected cognitive impairment, participants complete annual neuropsychological and neurologic evaluations until they develop dementia or are adjudicated to be cognitively normal. Assessments suggestive of possible mild cognitive impairment or dementia are followed by referral to our study dementia review committee, comprising of at least a neurologist and neuropsychologist, who adjudicate dementia diagnosis. The diagnosis of dementia is based on the criteria on DSM-IV criteria^4^ and additionally requires that the participant survive for at least 6 months following the onset of symptoms.

**iv. MrOS:**

The Osteoporotic Fractures in Men (MrOS) Sleep Study is an ancillary study to MrOS, a prospective cohort of men aged 65 and older, initially recruited from six clinical centers throughout the US from 2000 through 2002^6, 7^ [https://mrosonline.ucsf.edu](https://mrosonline.ucsf.edu/). The MrOS sleep study enrolled men aged 67 and older from 2003 through 2005. Incident dementia for MrOS participants during follow-up was defined as report of physician-diagnosed dementia, use of dementia medication (verified by clinic staff based on examination of pill bottles), or having a 3MS (Modified Mini-Mental State Examination) score<80. Follow up time for those with incident dementia was time to Sleep Visit 2 (November 2009 – March 2012) or time to Visit 4 (2014 -2016) since Sleep Visit 1 (December 2003 – March 2005) where PSG was measured, with a median follow-up time of 10.5 (Q1: 7.6, Q3: 11.4) years. Participants without dementia were censored at last contact data prior to or at Visit 4.

**v. SOF:**

The Study of Osteoporotic Fractures (SOF) study is a longitudinal cohort of women aged 65 and older, initially enrolled from 1986-1988 from 4 US clinical centers [https://sofonline.ucsf.edu](https://sofonline.ucsf.edu/). SOF participants were evaluated for dementia at Visit 9 (January 2007 – June 2008). Surveillance time was from baseline PSG (2002-2004) to Visit 9 (2007-2008), with a median follow-up time of 4.6 (Q1: 4.0, Q3: 5.2) years. Participants were identified as in need of cognitive adjudication (cases) when any one of the following criteria was met:

3MS Score of <88 (variable V93MSFLG)

CVLT Delayed (10 minute) Recall Score <4 (variable V9CVFLAG)

Functional Assessment Questionnaire (IQCODE) Score ≥3.6 (variable V9IQFLAG)

Self-reported history of dementia diagnosis (variable V9AZFLAG)

Living in a nursing home or personal care home (variable V9LVFLAG)

Those who met one or more of these criteria had current and past data forwarded to the adjudication committee (see full list of variables under the section heading “Data Used in the Adjudication Process”). This data was used by the adjudicators to determine the presence or absence of cognitive impairment. All adjudication was performed using existing data from the current and past visits. No further exam was performed on the participant, no additional data was gathered. If participants did not attend Visit 9, they were censored at the time of last contact prior to the visit. The participants who did not meet any of these criteria were considered cognitively normal and were not adjudicated.

**R code for Meta-analysis of 5 cohorts:**

Sleep exposures: N1%, N2%, N3%, REM%, SME%, WASO and TST.

library(meta)

library(readxl)

dat <- read_excel('Sleep_Meta_IncidentDementia_publish.xlsx')

dat1 <- dat[which((dat$Variable=="Percentage of stage 1 sleep, % of TST" |

dat$Variable=="Percentage of stage 2 sleep, % of TST" |

dat$Variable=="Percentage of slow wave sleep, % of TST" |

dat$Variable=="Percentage of REM sleep, % of TST")

& dat$Outcome=="Incident Dementia" & dat$Model==2),]

meta_id1 <- metagen(log(dat1$HR),

lower = log(dat1$CIL),

upper = log(dat1$CIU),

studlab=dat1$Cohort,

n.e=dat1$N,

n.c=dat1$Events,

pval = dat1$pvalue,

comb.fixed = FALSE,

comb.random = TRUE,

method.tau = "SJ",

adhoc.hakn.ci = "se",

adhoc.hakn.pi = "se",

prediction = FALSE,

sm = "HR",

byvar = dat1$Variable)

meta_id1

meta_id1$p.value <- meta:::formatPT(round(meta_id1$pval, 3),digits = 3)

randp1 <- meta:::formatPT(round(meta_id1$pval.random.w, 3),digits = 3)

dat2 <- dat[which((dat$Variable=="Sleep Maintenance Efficiency, %" |

dat$Variable=="Wake After Sleep Onset, min" |

dat$Variable=="Total sleep time: <=6 hours vs >6 (ref)")

& dat$Outcome=="Incident Dementia" & dat$Model==2),]

meta_id2 <- metagen(log(dat2$HR),

lower = log(dat2$CIL),

upper = log(dat2$CIU),

studlab=dat2$Cohort,

n.e=dat2$N,

n.c=dat2$Events,

pval = dat2$pvalue,

comb.fixed = FALSE,

comb.random = TRUE,

method.tau = "SJ",

adhoc.hakn.ci = "se",

adhoc.hakn.pi = "se",

prediction = FALSE,

sm = "HR",

byvar = dat2$Variable)

meta_id2

meta_id2$p.value <- meta:::formatPT(round(meta_id2$pval, 3),digits = 3)

randp2 <- meta:::formatPT(round(meta_id2$pval.random.w, 3),digits = 3)

#dev.off()

pdf(file="ID_Sleep_Meta_All_draft1.pdf",width=15,height=10)

plot.new()

plot.window(c(0,10),c(0,100))

#par(mar = c(1, 1, 1, 1))

meta::forest.meta(meta_id1,

layout='meta',

#plotwidth = "6cm",

#spacing = 0.75,

smlab=' ',

leftcols = c('studlab','n.c','n.e','effect', 'ci'),

leftlabs = c('Study','N Events','N','HR','95%CI'),

rightcols = c('p.value'),

rightlabs = c('P value'),

xlab='Incident Dementia',

#xlim=c(0.5,2),

ff.xlab='bold',

prediction=F,

col.diamond='lightblue',

col.by = "black",

col.square = "black",

col.inside = "black",

col.square.lines = "black",

subgroup.name = "",

print.subgroup.name = TRUE,

print.subgroup.labels = TRUE,

text.random.w = 'Pooled effects',

text.overall.random = '',

print.Q.subgroup = FALSE,

print.I2.ci = TRUE,

print.tau2 = FALSE,

print.Q = FALSE,

print.stat = FALSE,

test.subgroup.random = FALSE,

overall = FALSE,

overall.hetstat = FALSE

)

text(x=7.34,y=75,randp1[1],pos=4,font=2,col=1)

text(x=7.34,y=51,randp1[2],pos=4,font=2,col=1)

text(x=7.34,y=27,randp1[3],pos=4,font=2,col=1)

text(x=7.34,y=3.2,randp1[4],pos=4,font=2,col=1)

plot.new()

plot.window(c(0,10),c(0,100))

#par(mar = c(1, 1, 1, 1))

meta::forest.meta(meta_id2,

layout='meta',

#plotwidth = "6cm",

#spacing = 0.75,

smlab=' ',

leftcols = c('studlab','n.c','n.e','effect', 'ci'),

leftlabs = c('Study','N Events','N','HR','95%CI'),

rightcols = c('p.value'),

rightlabs = c('P value'),

xlab='Incident Dementia',

#xlim=c(0.5,2),

ff.xlab='bold',

prediction=F,

col.diamond='lightblue',

col.by = "black",

col.square = "black",

col.inside = "black",

col.square.lines = "black",

subgroup.name = "",

print.subgroup.name = TRUE,

print.subgroup.labels = TRUE,

text.random.w = 'Pooled effects',

text.overall.random = '',

print.Q.subgroup = FALSE,

print.I2.ci = TRUE,

print.tau2 = FALSE,

print.Q = FALSE,

print.stat = FALSE,

test.subgroup.random = FALSE,

overall = FALSE,

overall.hetstat = FALSE

)

text(x=7.34,y=63,randp2[1],pos=4,font=2,col=1)

text(x=7.34,y=39,randp2[2],pos=4,font=2,col=1)

text(x=7.34,y=15.5,randp2[3],pos=4,font=2,col=1)

dev.off()

**R code for meta-analysis of 3 Sleep Heart Health Study (SHHS) cohorts (secondary analysis):**

#Sleep exposures: N1%, N2%, N3%, REM%, SME%, WASO and TST.

library(meta)

library(readxl)

dat <- read_excel('Sleep_Meta_IncidentDementia_publish.xlsx')

dat1 <- dat[which((dat$Variable=="Percentage of stage 1 sleep, % of TST" |

dat$Variable=="Percentage of stage 2 sleep, % of TST" |

dat$Variable=="Percentage of slow wave sleep, % of TST" |

dat$Variable=="Percentage of REM sleep, % of TST")

& dat$Outcome=="Incident Dementia" & dat$Model==2

& dat$Study < 8),]

meta_id1 <- metagen(log(dat1$HR),

lower = log(dat1$CIL),

upper = log(dat1$CIU),

studlab=dat1$Cohort,

n.e=dat1$N,

n.c=dat1$Events,

pval = dat1$pvalue,

comb.fixed = FALSE,

comb.random = TRUE,

method.tau = "SJ",

adhoc.hakn.ci = "se",

adhoc.hakn.pi = "se",

prediction = FALSE,

sm = "HR",

byvar = dat1$Variable)

meta_id1

meta_id1$p.value <- meta:::formatPT(round(meta_id1$pval, 3),digits = 3)

randp1 <- meta:::formatPT(round(meta_id1$pval.random.w, 3),digits = 3)

dat2 <- dat[which((dat$Variable=="Sleep Maintenance Efficiency, %" |

dat$Variable=="Wake After Sleep Onset, min" |

dat$Variable=="Total sleep time: <=6 hours vs >6 (ref)")

& dat$Outcome=="Incident Dementia" & dat$Model==2

& dat$Study < 8),]

meta_id2 <- metagen(log(dat2$HR),

lower = log(dat2$CIL),

upper = log(dat2$CIU),

studlab=dat2$Cohort,

n.e=dat2$N,

n.c=dat2$Events,

pval = dat2$pvalue,

comb.fixed = FALSE,

comb.random = TRUE,

method.tau = "SJ",

adhoc.hakn.ci = "se",

adhoc.hakn.pi = "se",

prediction = FALSE,

sm = "HR",

byvar = dat2$Variable)

meta_id2

meta_id2$p.value <- meta:::formatPT(round(meta_id2$pval, 3),digits = 3)

randp2 <- meta:::formatPT(round(meta_id2$pval.random.w, 3),digits = 3)

#dev.off()

pdf(file="ID_Sleep_Meta_SHHS_draft1.pdf",width=15,height=10)

plot.new()

plot.window(c(0,10),c(0,100))

#par(mar = c(1, 1, 1, 1))

meta::forest.meta(meta_id1,

layout='meta',

#plotwidth = "6cm",

#spacing = 0.75,

smlab=' ',

leftcols = c('studlab','n.c','n.e','effect', 'ci'),

leftlabs = c('Study','N Events','N','HR','95%CI'),

rightcols = c('p.value'),

rightlabs = c('P value'),

xlab='Incident Dementia',

#xlim=c(0.5,2),

ff.xlab='bold',

prediction=F,

col.diamond='lightblue',

col.by = "black",

col.square = "black",

col.inside = "black",

col.square.lines = "black",

subgroup.name = "",

print.subgroup.name = TRUE,

print.subgroup.labels = TRUE,

text.random.w = 'Pooled effects',

text.overall.random = '',

print.Q.subgroup = FALSE,

print.I2.ci = TRUE,

print.tau2 = FALSE,

print.Q = FALSE,

print.stat = FALSE,

test.subgroup.random = FALSE,

overall = FALSE,

overall.hetstat = FALSE

)

text(x=7.31,y=69.5,randp1[1],pos=4,font=2,col=1)

text(x=7.31,y=51,randp1[2],pos=4,font=2,col=1)

text(x=7.31,y=32.5,randp1[3],pos=4,font=2,col=1)

text(x=7.31,y=14,randp1[4],pos=4,font=2,col=1)

plot.new()

plot.window(c(0,10),c(0,100))

#par(mar = c(1, 1, 1, 1))

meta::forest.meta(meta_id2,

layout='meta',

#plotwidth = "6cm",

#spacing = 0.75,

smlab=' ',

leftcols = c('studlab','n.c','n.e','effect', 'ci'),

leftlabs = c('Study','N Events','N','HR','95%CI'),

rightcols = c('p.value'),

rightlabs = c('P value'),

xlab='Incident Dementia',

#xlim=c(0.5,2),

ff.xlab='bold',

prediction=F,

col.diamond='lightblue',

col.by = "black",

col.square = "black",

col.inside = "black",

col.square.lines = "black",

subgroup.name = "",

print.subgroup.name = TRUE,

print.subgroup.labels = TRUE,

text.random.w = 'Pooled effects',

text.overall.random = '',

print.Q.subgroup = FALSE,

print.I2.ci = TRUE,

print.tau2 = FALSE,

print.Q = FALSE,

print.stat = FALSE,

test.subgroup.random = FALSE,

overall = FALSE,

overall.hetstat = FALSE

)

text(x=7.31,y=60.5,randp2[1],pos=4,font=2,col=1)

text(x=7.31,y=41.8,randp2[2],pos=4,font=2,col=1)

text(x=7.31,y=23.3,randp2[3],pos=4,font=2,col=1)

dev.off()

**Supplementary Table 1: Analytical sample selection**

|  | **ARIC** | **CHS** | **FHS** | **MrOS** | **SOF** |
| --- | --- | --- | --- | --- | --- |
| Participants with PSG and dementia surveillance, n | 1915 | 1229 | 936 | 2911 | 461 |
| Exclusions, n |  |  |  |  |  |
| <180 minutes of Total Sleep Time or <1 minute of REM sleep | 50 | 82 | 17 | 95 | 21 |
| Younger than 60 years | 680 | 0 | 541 | 0 | 0 |
| Prevalent dementia | 3 | 81 | 0 | 140 | 4 |
| Other neurological condition(s) | 28 | 72 | 8 | 96 | 58 |
| Missing covariates | 62 | 81 | 7 | 493 | 176 |
| Missing covariates sensitivity analysis additionally adjusting for apnea hypopnea index | 115 | 117 | 28 | 494 | 0 |
| **Final analysis sample** | **1092** | **913** | **363** | **2087** | **202** |
| **Final analysis sample for sensitivity analysis additionally adjusting for apnea hypopnea index** | **977** | **796** | **342** | **2086** | **202** |

ARIC, Atherosclerosis Risk in Communities study; CHS, Cardiovascular Health Study; FHS, Framingham Heart Study; MrOS, Osteoporotic Fractures in Men Study; SOF, Study of Osteoporotic Fractures; REM, rapid eye movement sleep; PSG, polysomnography. Model 1 adjusted for the effects of age (years) and sex (male, female). Model 2 included model 1 covariates and additionally adjusted for BMI (kg/m^2^), anti-depressant use (yes vs no), sedative use (yes vs no), and APOE e4 status (non ε4 carrier vs. at least one copy of ε4). Model 3 included model 2 adjustments with the addition of the apnea hypopnea index (AHI).

**Supplementary Table 2: Interaction (sleep macro-architecture x *APOE e4* status) P values: dementia outcome**

|  | **Sleep exposure x APOE e4 status**  **p value** | | | |
| --- | --- | --- | --- | --- |
|  | **ARIC** | **CHS** | **FHS** | **MrOS** |
| N1% | 0.873 | 0.155 | 0.684 | **0.026** |
| N2% | 0.354 | 0.126 | 0.997 | 0.711 |
| N3% | 0.749 | **0.016** | 0.775 | 0.706 |
| REM% | 0.339 | **0.031** | 0.523 | 0.143 |
| SME (%) | 0.272 | 0.773 | 0.868 | 0.425 |
| WASO (min) | 0.141 | 0.808 | 0.846 | 0.357 |
| Total sleep time (<6 vs ≥ 6 h) | 0.087 | 0.058 | 0.444 | 0.648 |

P values from Cox proportional hazard models for sleep exposure x APOE e4 status interaction for the outcome of incident dementia. All results are adjusted for age and sex. ARIC, Atherosclerosis Risk in Communities study; CHS, Cardiovascular Health Study; FHS, Framingham Heart Study; MrOS, Osteoporotic Fractures in Men Study; SOF, Study of Osteoporotic Fractures; N1, stage 1 non-rapid eye movement sleep; N2, stage 2 non-rapid eye movement sleep; N3, stage 3 non-rapid eye movement sleep; REM, rapid eye movement sleep; WASO, Wake after sleep onset; SME, sleep maintenance efficiency. *Stratified results for significant interactions are presented in Supplementary Table 5.*

**Supplementary Table 3: Association between sleep exposure and dementia incidence stratified by APOE e4 status**

| HR (95%CI), p value | | | | |
| --- | --- | --- | --- | --- |
|  | **CHS**  *APOE e4+* | *APOE e4-* | **MrOS**  *APOE e4+* | *APOE e4-* |
| N1% | - | - | 1.05 (1.01, 1.10), **0.027** | 0.98 (0.94, 1.03), 0.426 |
| N3% | 0.95 (0.86,1.05), 0.301 | 1.08 (1.00, 1.15), **0.045** | - | - |
| REM% | 1.02 (0.99,1.04), 0.197 | 0.98 (0.97,1.00), 0.053 | - | - |

Adjusted hazard ratios (HR) and 95% confidence intervals (95% CI) for incident dementia stratified by *APOE e4* status (*APOE e4+* and *APOE e4-)* in the Cardiovascular Health Study (CHS) and Osteoporotic Fractures in Men Study (MrOS) cohorts. Only those sleep exposure and dementia associations that were significantly moderated by APOE e4 status are presented. Data were adjusted for age and sex. N1, stage 1 non-rapid eye movement sleep; N3, stage 3 non-rapid eye movement sleep; REM, rapid eye movement sleep. Statistical significance, p<0.05.

**Supplementary Table 4: Interaction (sleep macro-architecture x sex) P values: dementia outcome**

|  | **Sleep exposure x sex p value** | | |
| --- | --- | --- | --- |
|  | **ARIC** | **CHS** | **FHS** |
| N1% | 0.080 | 0.951 | 0.330 |
| N2% | 0.953 | 0.123 | 0.063 |
| N3% | 0.238 | 0.201 | 0.383 |
| REM% | 0.616 | 0.554 | 0.090 |
| SME (%) | **0.049** | 0.933 | **0.017** |
| WASO (min) | 0.052 | 0.846 | **0.042** |
| Total sleep time (<6 vs ≥ 6 h) | 0.334 | 0.696 | 0.089 |

P values from Cox proportional hazard models for sleep exposure x sex interaction for the outcome of incident dementia. All results are adjusted for age. N1, stage 1 non-rapid eye movement sleep; N2, stage 2 non-rapid eye movement sleep; N3, stage 3 non-rapid eye movement sleep; REM, rapid eye movement sleep; WASO, wake after sleep onset; SME, sleep maintenance efficiency. *Stratified results for significant interactions are presented in Supplementary Table 5.*

**Supplementary Table 5: Association between sleep exposure and dementia incidence stratified by sex**

| HR (95% CI), p value | | | | |
| --- | --- | --- | --- | --- |
|  | **ARIC** |  | **FHS** |  |
|  | Women | Men | Women | Men |
| SME% | 1.09 (0.81, 1.46), 0.558 | 0.73 (0.53, 1.00), **0.047** | 0.45 (0.25, 0.80), **0.007** | 1.36 (0.68, 2.72), 0.388 |
| WASO (min) |  |  | 1.82 (1.08, 3.05), **0.024** | 0.79 (0.43, 1.43), 0.429 |

Adjusted hazard ratios (HR) and 95% confidence intervals (95% CI) for incident dementia stratified by sex (women and men) in the Atherosclerotic Risk in the Community (ARIC) and Framingham Heart Study (FHS) cohorts. Only those sleep and dementia associations that were significantly moderated by sex are presented. Wake after sleep onset (WASO); sleep maintenance efficiency (SME%). Statistical significance, p<0.05.

**Supplementary Table 6: Sensitivity analysis - Pooled Associations Between Each Sleep Measure and Dementia incidence**

| **Cohort** | **N** | **HR** | **Lower 95% CI** | **Upper 95% CI** | **p value** |
| --- | --- | --- | --- | --- | --- |
| **N1%** | | | | | |
| ARIC | 977 | 1.00 | 0.84 | 1.19 | 0.993 |
| CHS | 796 | 0.90 | 0.80 | 1.01 | 0.073 |
| FHS | 342 | 1.29 | 0.94 | 1.78 | 0.121 |
| MrOS | 2086 | 1.01 | 0.98 | 1.05 | 0.420 |
| SOF | 196 | 1.00 | 0.92 | 1.08 | 0.921 |
| Pooled effects, I^2^ 35% [0%; 76%] | 4397 | 1.00 | 0.91 | 1.10 | 0.965 |
| **N2%** | | | | | |
| ARIC | 977 | **0.99** | **0.98** | **1.00** | **0.048** |
| CHS | 796 | 1.00 | 0.99 | 1.00 | 0.181 |
| FHS | 342 | 1.00 | 0.98 | 1.02 | 0.913 |
| MrOS | 2086 | 1.01 | 0.98 | 1.03 | 0.669 |
| SOF | 202 | 0.99 | 0.95 | 1.02 | 0.457 |
| Pooled effects, I^2^ 0% [0.0%; 79%] | 4403 | 0.99 | 0.99 | 1.00 | 0.149 |
| **N3%** | | | | | |
| ARIC | 977 | **1.12** | **1.02** | **1.22** | **0.018** |
| CHS | 796 | 1.04 | 0.98 | 1.11 | 0.199 |
| FHS | 342 | 1.07 | 0.88 | 1.29 | 0.491 |
| MrOS | 2086 | 1.00 | 0.99 | 1.02 | 0.754 |
| SOF | 196 | 1.02 | 0.99 | 1.05 | 0.239 |
| Pooled effects, I^2^ 42% [0.0%; 79%] | 4397 | 1.03 | 0.99 | 1.07 | 0.117 |
| **REM%** | | | | | |
| ARIC | 977 | 0.99 | 0.97 | 1.01 | 0.386 |
| CHS | 796 | 1.01 | 0.99 | 1.02 | 0.490 |
| FHS | 342 | **0.94** | **0.90** | **0.98** | **0.007** |
| MrOS | 2086 | 0.98 | 0.95 | 1.02 | 0.262 |
| SOF | 202 | 0.98 | 0.91 | 1.05 | 0.570 |
| Pooled effects, I^2^ 56% [0.0%; 84%] | 4403 | 0.99 | 0.96 | 1.01 | 0.181 |
| **SME%** | | | | | |
| ARIC | 977 | 0.94 | 0.76 | 1.16 | 0.565 |
| CHS | 796 | 1.09 | 0.92 | 1.28 | 0.316 |
| FHS | 342 | 0.65 | 0.41 | 1.04 | 0.074 |
| MrOS | 2080 | 0.89 | 0.55 | 1.45 | 0.644 |
| SOF | 196 | 0.55 | 0.22 | 1.38 | 0.203 |
| Pooled effects, I^2^ 36% [0.0%; 76%] | 4391 | 0.90 | 0.71 | 1.13 | 0.369 |
| **WASO (min)** | | | | | |
| ARIC | 977 | 1.07 | 0.89 | 1.28 | 0.484 |
| CHS | 796 | 0.90 | 0.79 | 1.04 | 0.147 |
| FHS | 342 | 1.37 | 0.91 | 2.07 | 0.133 |
| MrOS | 2086 | 1.03 | 0.70 | 1.53 | 0.881 |
| SOF | 196 | **2.54** | **1.18** | **5.44** | **0.017** |
| Pooled effects, I^2^ 62% [0%; 86%] | 4397 | 1.17 | 0.85 | 1.60 | 0.344 |
| **Total sleep time (**<6 vs ≥ 6 h**)** | | | | | |
| ARIC | 977 | 1.00 | 0.78 | 1.29 | 0.994 |
| CHS | 796 | 1.09 | 0.90 | 2.31 | 0.374 |
| FHS | 342 | 1.21 | 0.73 | 2.00 | 0.459 |
| MrOS | 2086 | 1.14 | 0.72 | 1.80 | 0.574 |
| SOF | 202 | 0.70 | 0.28 | 1.75 | 0.447 |
| Pooled effects, I^2^ 36% [0%; 76%] | 4399 | 0.88 | 0.64 | 1.21 | 0.434 |

Adjusted hazard ratios (HR) and 95% confidence intervals (95% CI) for incident dementia. ARIC, Atherosclerosis Risk in Communities study; CHS, Cardiovascular Health Study; FHS, Framingham Heart Study; The Osteoporotic Fractures in Men, MrOS; The Study of Osteoporotic Fractures, SOF; N1, stage N1 sleep; N2, stage N2 sleep; N3, stage 3; REM, rapid eye movement; SME, sleep maintenance efficiency; WASO, wake after sleep onset.

**REFERENCES:**

1.	Knopman DS, Gottesman RF, Sharrett AR, Wruck LM, Windham BG, Coker L, et al. Mild cognitive impairment and dementia prevalence: The atherosclerosis risk in communities neurocognitive study (aric-ncs). *Alzheimers Dement (Amst)*. 2016;2:1-11

2.	Vahia VN. Diagnostic and statistical manual of mental disorders 5: A quick glance. *Indian journal of psychiatry*. 2013;55:220

3.	Fitzpatrick AL, Kuller LH, Ives DG, Lopez OL, Jagust W, Breitner JC, et al. Incidence and prevalence of dementia in the cardiovascular health study. *J Am Geriatr Soc*. 2004;52:195-204

4.	American Psychiatric Association. Diagnostic and statistical manual of mental disorders: Dsm-iv-tr. *American Psychiatric Pub*. 2000;157

5.	Satizabal CL, Beiser AS, Chouraki V, Chêne G, Dufouil C, Seshadri S. Incidence of dementia over three decades in the framingham heart study. *New England Journal of Medicine*. 2016;374:523-532

6.	Blank JB, Cawthon PM, Carrion-Petersen ML, Harper L, Johnson JP, Mitson E, et al. Overview of recruitment for the osteoporotic fractures in men study (mros). *Contemp Clin Trials*. 2005;26:557-568

7.	Orwoll E, Blank JB, Barrett-Connor E, Cauley J, Cummings S, Ensrud K, et al. Design and baseline characteristics of the osteoporotic fractures in men (mros) study--a large observational study of the determinants of fracture in older men. *Contemp Clin Trials*. 2005;26:569-585
